# Transmission onset distribution of COVID-19

**DOI:** 10.1101/2020.05.13.20101246

**Authors:** June Young Chun, Gyuseung Baek, Yongdai Kim

**Author notes:** Correspondence to: Yongdai Kim, Ph.D., Department of Statistics, Seoul National University, 56-1 Mountain, Sillim-dong, Gwanak-gu, Seoul, 08826, South Korea. June Young Chun and Gyuseung Baek contributed equally to this manuscript.

## Abstract

**Objectives:** The distribution of the transmission onset of COVID-19 relative to the symptom onset is a key parameter for infection control. It is often not easy to study the transmission onset time, as is difficult to know who infected whom exactly when.

**Methods:** We inferred transmission onset time from 72 infector-infectee pairs in South Korea, either with known or inferred contact dates by means of incubation period. Combining this data with known information of infector’s symptom onset, we could generate the transmission onset distribution of COVID-19, using Bayesian methods. Serial interval distribution could be automatically estimated from our data.

**Results:** We estimated the median transmission onset to be 1.31 days (standard deviation, 2.64 days) after symptom onset with peak at 0.72 days before symptom onset. The pre-symptomatic transmission proportion was 37% (95% credible interval [CI], 16–52%). The median incubation period was estimated to be 2.87 days (95% CI, 2.33–3.50 days) and the median serial interval to be 3.56 days (95% CI, 2.72–4.44 days).

**Conclusions:** Considering the transmission onset distribution peaked with the symptom onset and the pre-symptomatic transmission proportion is substantial, the usual preventive measure might be too late to prevent SARS-CoV-2 transmission.

## Introduction

SARS-CoV-2, a novel coronavirus first reported in Wuhan City, China in December 2019, has been spreading globally and the World Health Organization declared it as a pandemic on March 11, 2020 (Zhu et al., 2020). A total of 3,096,626 cases of coronavirus disease 2019 (COVID-19), and 217,896 confirmed deaths were counted world widely by April 30, 2020.

Considering its global threat, the transmission dynamics of COVID-19 should be made explicit. There have been studies reporting the estimated serial interval and incubation period, based on the early epidemics in China (Backer et al., 2020, Lauer et al., 2020, Li et al., 2020, Nishiura et al., 2020). The serial interval is the duration between the symptom onset of successive cases, and the incubation period is the time since infection to the symptom onset. Those are key epidemiological parameters, giving essential insight to infer the transmission potential or to determine the quarantine duration (White and Pagano, 2008).

There have been multiple reports of pre-symptomatic transmission of SARS-CoV-2, highlighting the difficulty of containment and mitigation of the disease (He et al., 2020, Liu et al., 2020, Wei et al., 2020). However, transmission onset distribution of COVID-19 has never been studied before. It is not easy to study the transmission onset time, as is difficult to know who infected whom exactly when. In this report, we tried to estimate the transmission onset distribution relative to the symptom onset by means of solid epidemiologic data of infector-infectee pairs in South Korea.

## Methods

### Data

We searched public reports of confirmed COVID-19 patients by government and each municipal website of South Korea. As of March 31, 9,887 cases have been confirmed of which 8,260 (83.5%) cases were linked to certain clusters such as religious groups, hospitals, or long-term healthcare facilities. Five-hundred and sixty cases (5.7%) were imported from abroad. We screened all available cases between January 23 to March 31, 2020 and selected the pairs with clearly defined contact history. We excluded cases from main clusters unless the causal relationship between an infector and infectee pair was evident.

For each pair of cases, we collected information on the dates of symptom onset of both the infector and the infectee, exposure dates of the infectee by the infector, and the dates of confirmation of both. Considering the South Korean quarantine policy, the confirmation date could be regarded as the date on which isolation started. Demographic information of age and sex were also gathered.

### Procedures

We tried to estimate the time difference between an infector’s symptom onset and its transmission onset time. We defined symptom onset date (*s*), an infectee’s infection date (*1*), an infector’s transmission onset date (*P*), and contact time of the pair as (*C*_*L*_, *C*_*R*_), where subscripts L and R denote the left and right boundaries, meaning the earliest and the latest contact dates respectively. The aim of the study is to estimate the timing of *P* relative to the *s* (*P-s*), which we denominate as *W*. We put as an infectee and *J*_*i*_ as the corresponding infector, and observed (*s*_*j*_i, *C*_*L,i*_, *C*_*R,i*_, *s*_*i*_). *1*_*i*_ should be within interval [*C*_*L,i*_, *C*_*R,i*_], and transmission onset of an infector (*P*_*j*_) is described as 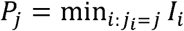. Using the pair *l*(*s*_*j*_, *P*_*j*_)| *J, nfector*, we were able to estimate the distribution *f*(*W*) of *W = P − s*.

There are three possible scenarios for *I*_*i*_ in our data.

i. There is an exact calendar date of an exposure. In this case, we could directly achieve the timing of transmission as *I*_*i*_ *= C*_*L,i*_ *= C*_*R,i*_.
ii. There is a specified duration of exposure (*C*_*L,ii*_< *C*_*R,i*_).
iii. There is a continuous exposure such as household members (*C*_*L,i*_ *= −* ∞, *C*_*R,i*_ *= s*_*i*_).

Recall that the aim is to estimate the distribution *f*(*W*) of *W* based on the data 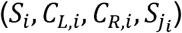 for infectees. We consider the following parametric model for *f*(*W*):

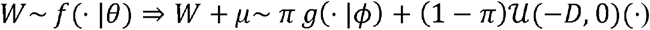

where *θ =* (*ϕ, µ, π*), *µ* > *0, π* ∈ (0,1), *g*(|*ϕ*) is the gamma distribution with the parameter *ϕ* and *‘*𝓊(*-D, 0*)(·) is the uniform distribution with the support (− *D, 0*). In the model, *µ* is the shift parameter for cases with transmission prior to the symptom onset. We introduced the term (1 − *π*)𝓊(− *D, 0*)(·) to make the effect of negative outliers of *W* be minimal to the estimation of the parameter. That is, we considered *W* to be an outlier if *W < - µ*. In the model, *D* > *0* is an arbitrary limit for an outlier of *W*. In the analysis, we used *D* = 14, since the usual incubation period is within 14 days.

A difficulty in estimating *θ* is that the exact *W* is not observed for cases in categories (ii) and (iii). However, conceptually we could impute the timing of transmission *I* by use of the estimated incubation period distribution (Reich et al., 2009). With our notation, *z = s* − *I* is the incubation period. From our data of contact dates and symptom onset dates, we could estimate the distribution of *z* which we utilized to impute *I*. That is, in case (ii), we could generate *z* from conditional probability distribution *z*| *z* ∈ [*s-C*_*R*_,*s* − *C*_*L*_], and then we let *I = s* − *z*. Similarly, we could generate *z* from conditional probability distribution *Z* |*Z* ≤ *S* for case (iii). To accommodate this idea, we used a Bayesian method and obtained the posterior distribution of *θ* based on the Markov chain Monte Carlo (MCMC) algorithm in which *W* is imputed for cases in categories (ii) and (iii). A detailed description of the MCMC algorithm is provided in Supplementary Material.

For sensitivity analysis, we utilized the incubation period distribution of a previous study and replicated the same analysis as above (Lauer et al., 2020). Moreover, we hold the parameter *µ* as 4, and compared the result using the same procedure, since the minimum of *W* from our data was -4 days. Serial interval distribution could be automatically estimated from our data.

All analyses were conducted using the *R* statistical software version 3.6.3. All code and data are available in the supplementary material. We present case numbers with randomization process, instead of nationally designated identification numbers, in order to maintain confidentiality.

## Ethics Approval

All the data used in this study were publicly available and were approved from institutional review board assessment of National Cancer Center (NCC2020-0119).

## Results

We found 89 infectees with defined source of infection (infector) and contact history. Four infectors (4.5%) were asymptomatic until diagnosed, and thus were excluded from the analysis. Sixteen infectees (18.0%) were asymptomatic when diagnosed, among which 13 cases had no specified contact date and were unable to guess the infection time. In short, 72 infector-infectee pairs were included in the study (Fig 1).

**Fig 1.**
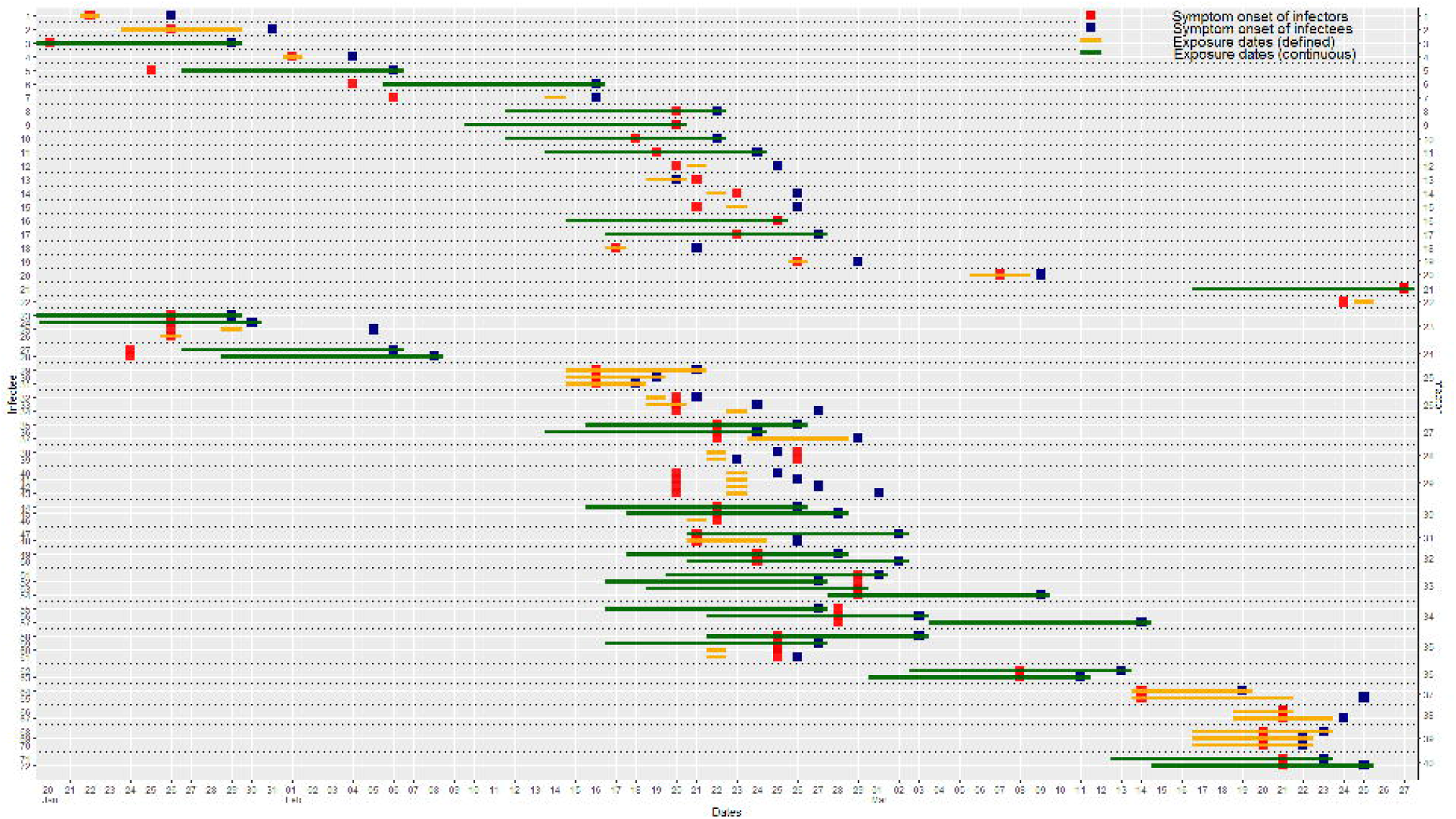
Transmission plot of infector-infectee pairs. For cases of continuous exposure, the interval presentation was limited to 10 days prior to the symptom onset whoever came first, considering the upper 95^th^ percentile of pre-estimated incubation period is 10.1.

Twenty-two cases documented a single point date of contact, and 16 cases had documented duration of contact dates. Otherwise, 34 cases (32 household members and 2 colleagues) had continuous exposure history (Supplementary Material). The median age of the 72 infectees was 40 years (interquartile range [IQR], 24 to 54 years). Of these, 34 (47%) were male and 38 (53%) were female. The infectees had 40 unique infectors. The average number of transmissions per infector was 1.8, with a maximum of four cases (Fig 2).

**Fig 2.**
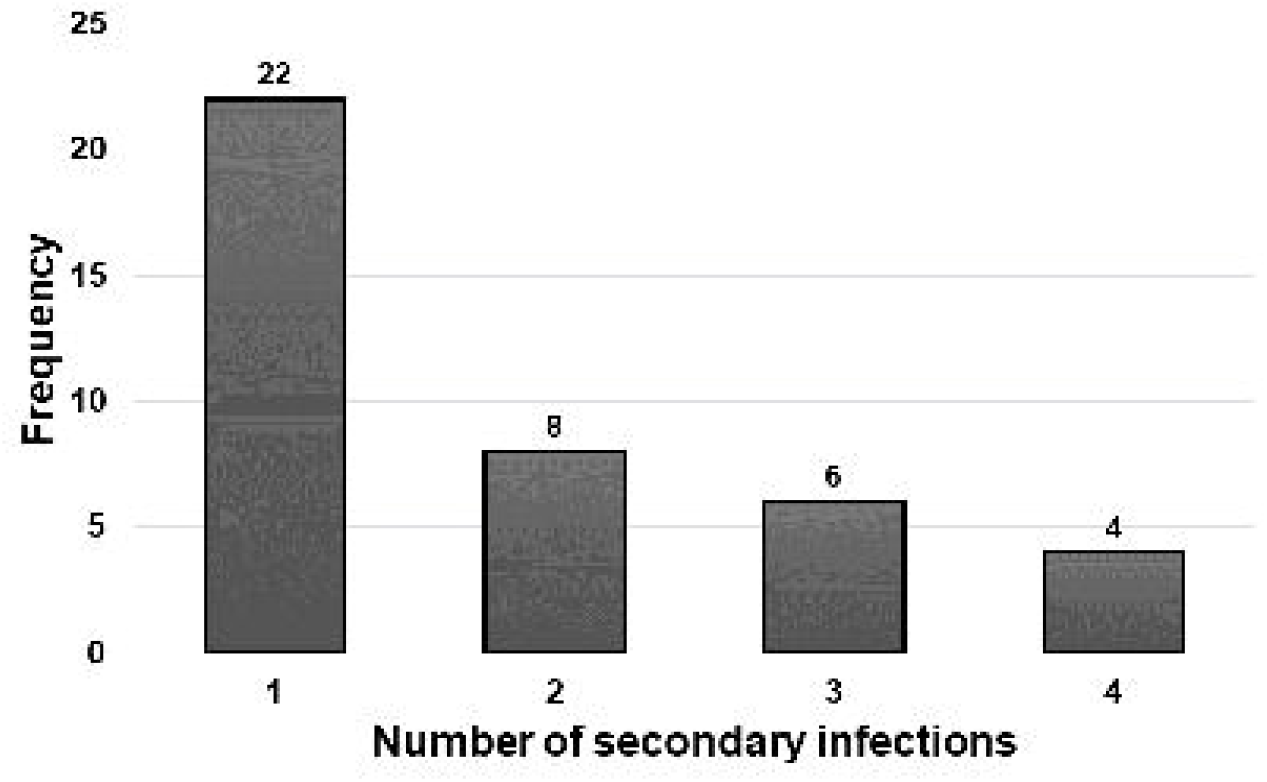
Number of secondary infections per infector in the reported data set.

We estimated the incubation period distribution of COVID-19 using a lognormal model. Among 35 pairs with given contact dates, the median incubation period was 2.87 days (95% credible interval [CI], 2.33–3.50) (Table 1). The estimated median serial interval from 69 infector-infectee pairs with given symptom onset dates was 3.56 days (95% CI, 2.72–4.44), with the best-fit using a lognormal distribution (Table 2).

**Table 1.** Estimated incubation period distributions for COVID-19 based on 35 infector-infectee pairs with defined contact dates.

|  | Lognormal | Gamma | Weibull |
| --- | --- | --- | --- |
| Median (95% CI) | 2.87 (2.33–3.50) | 2.99 (2.44–3.60) | 3.10 (2.54–3.71) |
| Parameter 1 | 1.055 (0.845–1.254) | 4.544 (2.590–10.494) | 2.310 (1.747–3.743) |
| Parameter 2 | 0.493 (0.301–0.643) | 0.709 (0.312–1.178) | 3.635 (3.030–4.208) |
| Log-likelihood | -46.6 | -45.4 | -45.1 |

For the lognormal distribution, parameter 1 and parameter 2 are the mean and SD of the natural logarithm and for all other distributions, parameter 1 and parameter 2 are the shape and scale parameters, respectively.

**Table 2.** Estimated serial interval distributions for COVID-19 based on 69 infector-infectee pairs with symptom onset dates.

|  | Lognormal | Gamma | Weibull |
| --- | --- | --- | --- |
| Median (95% CI) | 3.56 (2.72–4.44) | 3.75 (2.87–4.72) | 3.93 (3.04–4.92) |
| Parameter 1 | 3.18 (2.22–4.24) | 3.45 (2.56–5.19) | 1.96 (1.64–2.44) |
| Parameter 2 | 0.75 (0.47–1.03) | 2.19 (1.44–2.91) | 8.47 (7.53–9.53) |
| Log-likelihood | -77.5 | -192.8 | -189.8 |

For the lognormal distribution, parameter 1 and parameter 2 are the mean and SD of the natural logarithm and for all other distributions, parameter 1 and parameter 2 are the shape and scale parameters, respectively.

Based on the 72 infecfor-infectee pairs, the Bayes estimate of the transmission onset distribution after a smoothing procedure was given as Fig 3A. The mean and median values were 1.31 days (95% CI, 0.38–2.55) and 0.68 days (95% CI, −0.09–1.73) after infectors’ symptom onset, respectively, with the peak at 0.72 days before symptom onset. The pre-symptomatic transmission proportion was 37% (95% CI, 16–52%).

**Fig 3.**
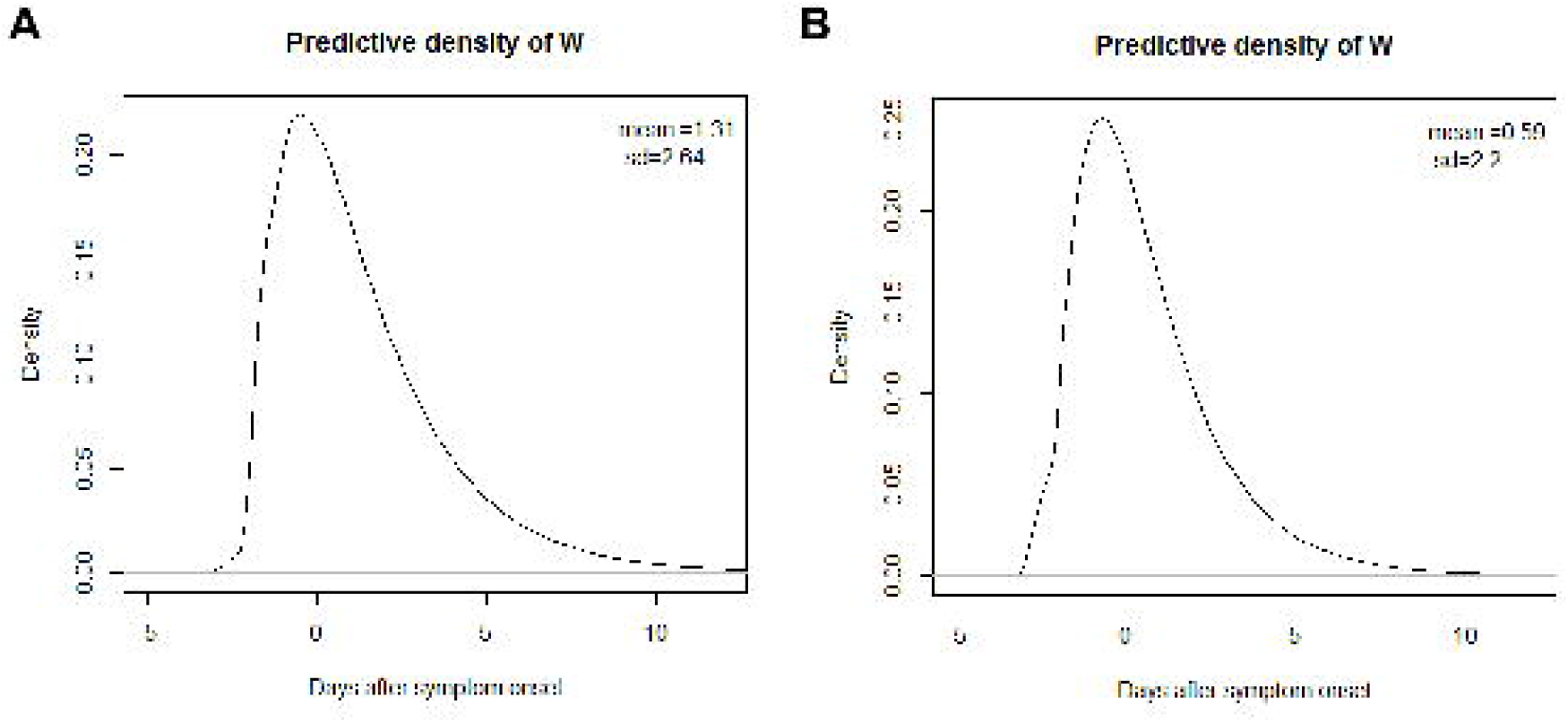
Estimated distribution (*W*) of the transmission onset of COVID-19 relative to the symptom onset (A) using the incubation period distribution in the current study, and (B) using the previous incubation period distribution. (Lauer et al., 2020)

For sensitivity analysis, applying the incubation period of previous study, the transmission onset distribution was inferred as Fig 3B. The mean and median values were 0.59 days (95% CI, −0.33–1.79) and 0.12 days (95% CI, −0.75–1.06) after the infector’s symptom onset, respectively, with the peak at 0.41 days before symptom onset. The pre-symptomatic transmission proportion was 48% (95% CI, 29–67%). With a fixed parameter of *µ* as 4, the mean and median were 0.59 (95% CI, −0.34–1.79) and −0.07 days (95% CI, −0.91–0.99), with the pre-symptomatic transmission proportion of 51% (95% CI, 37–64%).

## Discussion

The transmission onset distribution is of utmost interest to clinicians and public health workers, but it has not been widely studied to date due to lack of epidemiological information. Based on concrete data, we hereby present that the transmission of COVID-19 could start before the symptom onset, and the probability peaked as the symptom start and declined thereafter. Interestingly, the trend of this distribution looks similar with published SARS-CoV-2 viral load kinetics though it could not include the viral load data before the symptom onset (Kim et al., 2020, Zou et al., 2020).

In South Korea, the first COVID-19 case was identified on January 19, 2020 in a traveler from Wuhan City, who was quarantined immediately at the airport screening. Until February 18, 2020, there were only imported cases from abroad or cases from their close contacts who were under the surveillance. The situation was totally changed after an outbreak with an unknown source of infection occurred within a religious community, resulting in a total of 5,210 confirmed cases in that single cluster as of April 12, 2020, a number which is almost half (49.6%) of the total 10,512 cases in South Korea. This study spanned from late January to the end of March, 2020, and encompassed all the phases observed in South Korea, thus enhancing the reliability of the results. Furthermore, in South Korea close contacts of confirmed patients were screened extensively with SARS-CoV-2 nucleic acid testing, regardless of whether the contact had symptoms. Given this situation, it can be assumed that cases of infection are unlikely to have been missed.

Among the 89 infectors, 4 cases (4.5%) were asymptomatic when diagnosed, but out of 89 infectees, 16 cases (18.0%) were asymptomatic when diagnosed. The difference is attributed to the active surveillance of close contacts of COVID-19 patients, including the screening tests of those asymptomatic, which in turn, pre-symptomatic. Our study revealed that approximately 40% of individuals with COVID-19 infected others when they could not acknowledge any symptom. This finding is in line with other recently published studies revealing pre-symptomatic transmission ratio of 46% (95% CI, 21-46%), 44% (95% CI, 25-69%) in China, and 48% (95% CI, 32-67%) in Singapore (Ganyani et al., 2020, He et al., 2020, Liu et al., 2020).

This study has several limitations. First, the dates of symptom onset and contact time were identified from epidemiological investigation and it could be false due to recall bias. Second, we only collected data from definite infector-infectee pairs with defined contact times. Hence, our estimation of transmission onset distribution is the upper bound of the true value. Third, considering the substantial proportion of pre-symptomatic transmission, it is possible that one infectee had multiple infectors, and so-called effective contacts could not be identified. Fourth, we excluded clustered data to promote the accuracy of causal relationships. The transmission behavior of SARS-CoV-2 might have different characteristics within unique clusters, but this could not be measured in this study. Fifth, we might not catch the late-onset transmission cases, considering the pre-emptive screening and isolation measures implemented in South Korea.

This study provides an important message in terms of public health practice. It indicates that the usual preventive measure of isolating people when they become symptomatic, might be too late to prevent SARS-CoV-2 transmission.

## Data Availability

All code and data are available in the supplementary material.

## Data availability

All code and data are available in the supplementary materials. We present case numbers with randomization process, instead of nationally designated identification numbers, in order to maintain confidentiality.

## Author contributions

J.Y.C. and Y.K. conceived of the study, and G.B. performed the analysis. J.Y.C. and G.B. wrote the first draft of the manuscript. Y.K. reviewed and edited the manuscript. All authors interpreted the findings, contributed to writing the manuscript, and approved the final version for publication.

## Acknowledgement

We acknowledge public health authorities in South Korea for their innumerable efforts to perform epidemiologic investigation to stop the COVID-19 outbreak.

